# Gene expression has distinct associations with brain structure and function in major depressive disorder

**DOI:** 10.1101/2022.09.20.22280083

**Authors:** Shu Liu, the DIRECT Consortium, Abdel Abdellaoui, Karin J.H. Verweij, Guido A. van Wingen

## Abstract

Major depressive disorder (MDD) is associated with structural and functional brain abnormalities. MDD as well as brain anatomy and function are influenced by genetic factors, but the role of gene expression remains unclear. Here we investigated how cortical gene expression contributes to structural and functional brain abnormalities in MDD. We compared the gray matter volume and resting-state functional measures in a Chinese sample of 848 MDD patients and 749 healthy controls, and we then associated these case-control differences with cortical variation of gene expression. While whole gene expression was positively associated with structural abnormalities, it was negatively associated with functional abnormalities. We observed the relationships of expression levels with brain abnormalities for individual genes, and found that transcriptional correlates of brain structure and function showed opposite relations with gene dysregulation in postmortem cortical tissue from MDD patients. We further identified genes that were positively or negatively related to structural abnormalities as well as functional abnormalities. The MDD-related genes were enriched for brain tissue, cortical cells, and biological pathways. These findings suggest that distinct genetic mechanisms underlie structural and functional brain abnormalities in MDD, and highlight the importance of cortical gene expression for the development of cortical abnormalities.

## Introduction

Major depressive disorder (MDD) is a highly prevalent psychiatric disorder [1] that is the third leading cause of disability worldwide [2]. Despite decades of research, the pathophysiology of MDD is not well understood. The etiology of MDD is complex and influenced by genetic factors. Twin studies have estimated that the heritability of MDD is approximately 37% [3], and genome-wide association studies (GWASs) have captured ∼9% of the heritability with common single nucleotide polymorphisms (SNPs) [4]. Moreover, MDD is highly polygenic, resulting from the joint effects of many genetic variants with small effects scattered across the genome [5].

Previous research has shown that certain brain regions are affected in MDD individuals. There are structural and functional differences between depressed individuals and healthy controls in various brain regions (e.g. the anterior cingulate and frontal regions) [6–10]. To investigate the neural mechanisms of genetic risk for MDD, neuroimaging intermediate phenotypes can be used to link genetic variation to this complex psychiatric syndrome [11]. For example, the genetic predisposition to MDD is associated with brain structure in the orbitofrontal cortex [12] and brain function in the lateral frontal and precentral regions [13]. Although imaging genetics analyses for MDD have provided evidence that genetic factors play a major role in these brain phenotypes implicated in depression, the intermediate processes remain to be elucidated.

Gene expression is the most fundamental molecular process through which genes can regulate the differentiation, development, and functioning of brain cells and tissues [14]. Expression levels of genome-wide genes have been estimated in post-mortem brain tissue from six donors, and have been made public as the Allen Human Brain Atlas (AHBA) [15] and this information has been widely used to examine the transcriptional correlates of brain structure and function [16]. Brain-wide gene expression data has been associated with cortical volume [17], MRI signal intensity [18], and resting-state activity [19, 20]. Previous studies that combined neuroimaging and AHBA gene expression data identified several genes related to structural brain abnormalities in various psychiatric disorders [21–23]. A previous study has linked depression-related cortical abnormalities to normative patterns of gene expression [24] in healthy individuals with negative affect measures and individuals who have experienced depressive illness in the past. Another study by Li et al [25] found a positive correlation between gene expression and structural brain abnormalities of morphometric similarity in patients with depression. However, this study used a relatively small sample size which could have led to insufficient statistical power to detect important associations, and the study only focused on structural brain abnormalities and did not consider functional brain abnormalities.

The aim of this study was to extend the work by Li et al. [25] by substantially increasing the sample size and by including brain function. We used T1 and resting-state functional MRI data from 848 MDD patients and 794 healthy controls to compare gray matter volume (GMV) and three major functional measures in 210 cortical brain regions based on Brainnetome (BN) atlas [26]. We then linked the expression level of 15,633 genes in these regions to the identified case-control structural and functional differences. The UK Biobank dataset was used to replicate the transcriptome-neuroimaging relationships. We further investigated the relationships of transcriptional correlates of in vivo brain abnormalities with ex vivo gene dysregulation for psychiatric disorders. Moreover, we selected genes transcriptionally involved in structural or functional brain differences, and performed gene set enrichment analysis for tissues, cortical cell types, and biological pathways to understand the functional annotations of identified gene sets.

## Methods and Materials

### Participants

In our main analysis, we used publicly available data with brain scans from the REST-meta-MDD project [27, 28]. The project totally included 1,300 patients diagnosed with MDD and 1,128 healthy controls. From them, 848 patients diagnosed with MDD (534 females and 314 males, age ranges: 18-65 years old) and 794 healthy controls (467 females and 327 males, age ranges: 18-64 years old) from 17 sites in China were extracted for group comparison analysis (**Figure S1**). Age and sex did not have difference between the MDD and control group (age: *t* = -0.75, *p* = 0.94, sex: χ^2^ = 2.8, *p* = 0.094). Individuals with MDD were diagnosed as having current depressive status by experienced psychiatric physicians based on ICD10 or DSM-IV. All participants provided written informed consent and the project was approved by local Institutional Review Boards (for details see **Table S1**).

The REST-meta-MDD dataset was comprised of east Asian individuals from China. To assess whether the results can be generalized to other ethnic populations, we replicated the analyses in a European-ancestry sample from the UK Biobank (802 depressed individuals and 802 controls) (**Figure S2, Table S2**), matched for age, sex, intelligence, and educational attainment. More information about the sample selection, and phenotypic measures in the UK Biobank sample can be found in **SI Methods**.

### Imaging procedures

Structural and functional MRI data were collected from 17 scanning sites using different 3T or 1.5T scanners, and different parameters were used for imaging acquisition (**Table S1**). A MATLAB- and SPM-based pipeline, DPARSF[29], was used to preprocess the resting-state functional MRI (rsfMRI) [27]. Main procedures included removing the first 10 time points, slice-timing correction, realignment, regressing out head motion effects using the Friston 24 parameter model [30], and normalization from individual native space to MNI space using DARTEL tool [31]. After realignment, individual T1 weighted images were co-registered to the mean functional image and then segmented into gray matter (GM) and white matter. GM images were also modulated and spatially normalized by the DARTEL tool.

After preprocessing, three main functional brain measures (amplitude of low-frequency fluctuation (ALFF), fractional ALFF (fALFF), and regional homogeneity (ReHo)) were calculated using rsfMRI data. Next, all functional images were Z standardized and then smoothed with a 3 mm full-width at half maximum (FWHM) Gaussian kernel. Similarly, GM images were also smoothed with the 3 mm FWHM Gaussian kernel. Finally, smoothed GM and three functional images were mapped to the BN atlas to extract GM volume (GMV), ALFF, fALFF and ReHo of 210 cortical regions, respectively. More details about the imaging acquisition and processing can be found in the study by Yan et al. [27]. The UK Biobank data were analyzed using a series of imaging processing steps which can be found in **SI Methods**.

### Estimation of regional gene expression

We used the AHBA microarray DNA data (http://human.brain-map.org), comprising gene expression measures from a total of 3702 tissue samples by probes[15]. The tissues were collected from six human donor brains 24 to 57 years of age with no known neuropsychiatric or neuropathological history (5 males and 1 female). Of the donors, there were three Caucasian, two African-American and one Hispanic. Then, the abagen toolbox [32] in Python was used to create a whole-genome expression atlas by probe-to-gene and sample-to-region strategies using the following steps: (1) intensity-based filtering of microarray probes to remove those that do not exceed background noise of 50%; (2) selection of a representative probe for each gene across each hemisphere; (3) matching of microarray samples to brain parcels from the BN atlas; and (4) normalization, and aggregation within parcels and across donors [16]. This resulted in a 2D matrix that showed gene expression levels of 15,633 genes on 210 BN cortical regions. In addition, another cortical parcellation with 68 cortical regions, the Desikan-Killiany (DK) atlas, was used to extract AHBA gene expression matrix for replication analysis.

### Statistical analysis

We used two-sample *t* tests to obtain the case-control differences in GMV, ALFF, fALFF and ReHo based on the BN atlas. T-scores were then converted to Cohen’s d effect sizes for ease of interpretation. The covariates, including age, sex, education, and head motion, were regressed out of the brain measures. The false discovery rate (FDR) correction was applied to correct for multiple comparisons using the Benjamini and Hochberg method with a threshold of *p*=0.05. For a given brain measure, Cohen’s d across 210 cortical regions was used to quantify structural or functional abnormalities in MDD. Principle component analysis (PCA) was used to extract the first principal component (PC) of effect sizes for the three functional measures, defined as funcPC1, that explained most variance of functional abnormalities. We also extracted the first PC from the gene expression matrix, defined as genePC1.

To test the relationship of whole gene expression of 15,633 genes for 210 cortical regions with the corresponding brain abnormalities, we examined the correlation of genePC1 with GMV differences and funcPC1 using the Spearman’s rank correlation analysis.

To replicate the whole transcriptome-neuroimaging relationships, we conducted identical analyses in the UK Biobank dataset (**SI Methods**). In addition, we examined relationships between cortical expression distribution of individual genes and brain abnormalities, indicating individual gene’s transcriptional correlates of MDD-related brain abnormalities. A Bonferroni multiple comparison correction was applied to select the genes significantly positively or negatively related to GMV differences, defined as GMV+ genes or GMV- genes. Similarly, genes related to funcPC1 were defined as funcPC1+ or funcPC1- genes.

### Differential gene expression analysis in major psychiatric disorders

Following the approach of Gandal et al.[33], we obtained the meta-analytic estimates of differential gene expression (DGE) in postmortem brain tissues of patients with major psychiatric disorders including MDD, schizophrenia (SCZ), autism spectrum disorder (ASD), and bipolar disorder (BP) (**Table S3**). Through linear mixed effects modeling, standardized beta coefficients were calculated as DGE values, which quantified the degree that a gene is up- or down-regulated for a given psychiatric population. The tissues were sampled mainly from the prefrontal and anterior cingulate cortex of MDD patients, and the prefrontal cortex only for other psychiatric patients.

We obtained the ∼10,000 overlapping genes between AHBA and Gandal et al. [33] datasets, and examined the associations between transcriptional correlates of brain abnormities in MDD and DGE values for psychiatric disorders for\ these common genes using Spearman’s rank correlation analysis. It has been suggested that linking transcriptional correlates to DGE values directly for all genes could cause the bias for estimating correlation because of the noise [24], we further applied the bin-based correlation analysis. In detail, the common genes were ranked by their correlations with brain abnormalities (transcriptional correlates) in descending order. We then divided the ranked genes into 100 bins, which merged the genes with similar transcriptional correlates of brain abnormalities into a bin. For example, the genes with highest positive and negative correlations were divided into the 1st and 100th gene bin, respectively. The average transcriptional correlates and DGE values were calculated for each bin. For 100 gene bins, we examined the relationships of average transcriptional correlates of in vivo brain abnormalities with ex vivo DGE values for psychiatric disorders. Alternative numbers of bins (N = 50, 150, and 200) were also applied to test the stability across choices of bin numbers.

### Enrichment analysis

We applied gene set enrichment analysis to identify functional annotations of our identified genes related to brain abnormalities in MDD (GMV+ genes, GMV- genes, funcPC1+ genes, and funcPC1- genes). First, we conducted tissue-specific expression analysis (TSEA)[34]. A specificity index probability (pSI = 0.05, permutation corrected) and Fisher’s exact test with FDR-BH correction were used to determine how likely our identified genes were to be specifically expressed in a given tissue. Next, we conducted cell-type expression analysis based on the study by Seidlitz et al. [35]. They provided gene sets related to seven canonical cortical cell classes: inhibitory neurons, excitatory neurons, astrocytes, microglia, endothelial cells, oligodendrocyte precursors, and oligodendrocytes (**Table S4**). We used the GSEApy package [36] in Python to identify the cell types that genes related to brain abnormalities were enriched for. Moreover, we selected the genes with the largest 300-800 positive and negative DGE values as up- and down-regulated gene sets for psychiatric disorders. Using the enrichment analysis, we tested whether up- and down-regulated genes for psychiatric disorders were expressed most in cortical regions that showed structural and functional abnormalities in MDD. In addition, the Enrichr tool available online (http://amp.pharm.mssm.edu/Enrichr) was used to identify enrichment pathways for the gene ontology (GO) biological processes and Kyoto Encyclopedia of Genes and Genomes (KEGG) pathways with FDR-BH correction [36].

## Results

### Case-control brain differences

Using the REST-meta-MDD dataset with 848 MDD patients and 794 healthy controls, we compared the case-control differences of three major functional brain measures (ALFF, fALFF, and ReHo) in 210 cortical regions. We found similar case-control difference patterns across the brain for these three functional measures, generally with increased neuronal activity in lateral regions and decreased activity in medial regions, although different statistically significant brain functional abnormalities were observed after FDR multiple testing correction (**Figure 1A, Figure S3**). MDD patients had significantly decreased ALFF in the left medial superior frontal cortex and significantly increased ALFF in inferior parietal cortex and temporal regions (**Figure S3A)**, whereas the MDD patients had significant decreased fALFF in the medial superior frontal and caudodorsal/pregenual cingulate cortex (**Figure S3B**). In MDD patients, ReHo was significantly increased in the lateral prefrontal cortex, and decreased in the dorsal insula, precentral/postcentral/paracentral cortex, and caudodorsal cingulate cortex (**Figure S3C**). The effect sizes for these functional differences ranged from Cohen’s d -0.2 to 0.2, Moreover, brain-wide case-control differences for ALFF, fALFF, and ReHo were highly positively correlated with each other (correlation *r*_*S*_ ranged from 0.43 to 0.71) (**Figure 1D, Figure S3D**). We therefore used PCA to extract the first PC of effect sizes for functional measures, defined as funcPC1, which explained 71% variance of functional brain abnormalities in MDD (**Figure 1B**).

**Figure 1.**
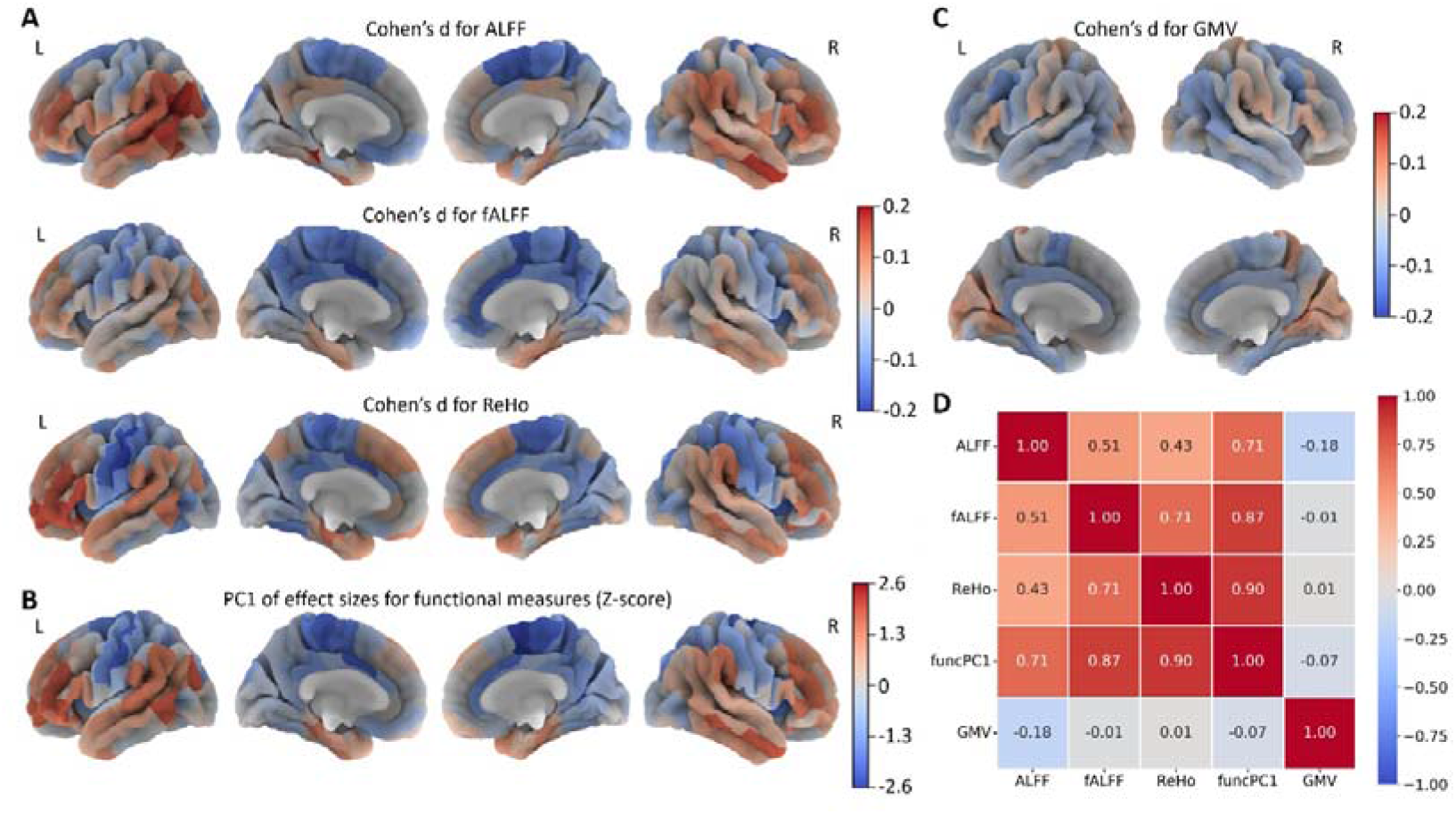
Case-control structural and functional brain differences in major depressive disorder (MDD)for 210 cortical regions. **(A)** Differences (Cohen’s d) for functional brain measures including amplitude of low-frequency fluctuation (ALFF), fractional ALFF (fALFF), and regional homogeneity (ReHo); **(B)** The first principle component of effect sizes for three functional measures (funcPC1) (Z-score normalization); **(C)** Differences (Cohen’s d) for gray matter volume (GMV); **(D)** The correlations between the case-control differences.

GMV showed smaller effect sizes (Cohen’s d: -0.1 - 0.12), with decreased GMV in frontal and temporal cortex and increased GMV in occipital and parietal regions (**Figure 1C**), but none of these case-control differences were significant.

To verify whether the observed case-control differences were dependent on the brain parcellation scheme, we repeated the analyses based on the Desikan-Killiany (DK) atlas with 68 cortical regions. These results showed a comparable pattern of structural and functional differences (**Figure S4**).

### Relationships of gene expression with case-control differences

PCA was also used to extract the first PC from the gene expression matrix (genePC1), which explained 18.6% of the variance (**Figure 2A**). We then estimated the association of genePC1 with funcPC1 as well as Cohen’s d for GMV and the functional measures. GenePC1 was negatively correlated withfuncPC1 (*r*_*S*_ = -0.281, *p* = 3.64 × 10^−5^) (**Figure 2B**), and genePC1 consistently showed negative correlation with effect sizes for ALFF, fALFF, and ReHo (**Figure S5**). In contrast, genePC1 was positively correlated with GMV differences (*r*_*S*_ = 0.367, *p* = 4.36 × 10^−8^) (**Figure 2C**). The opposite relationships of whole gene expression with funcPC1 and GMV differences indicates that the distribution of gene expression in the cortex is differentially related to functional and structural difference patterns in MDD. This pattern of opposite relationships of gene expression with structural and functional brain abnormities was replicated using data from the UK Biobank (**Figures S6-7**).

**Figure 2.**
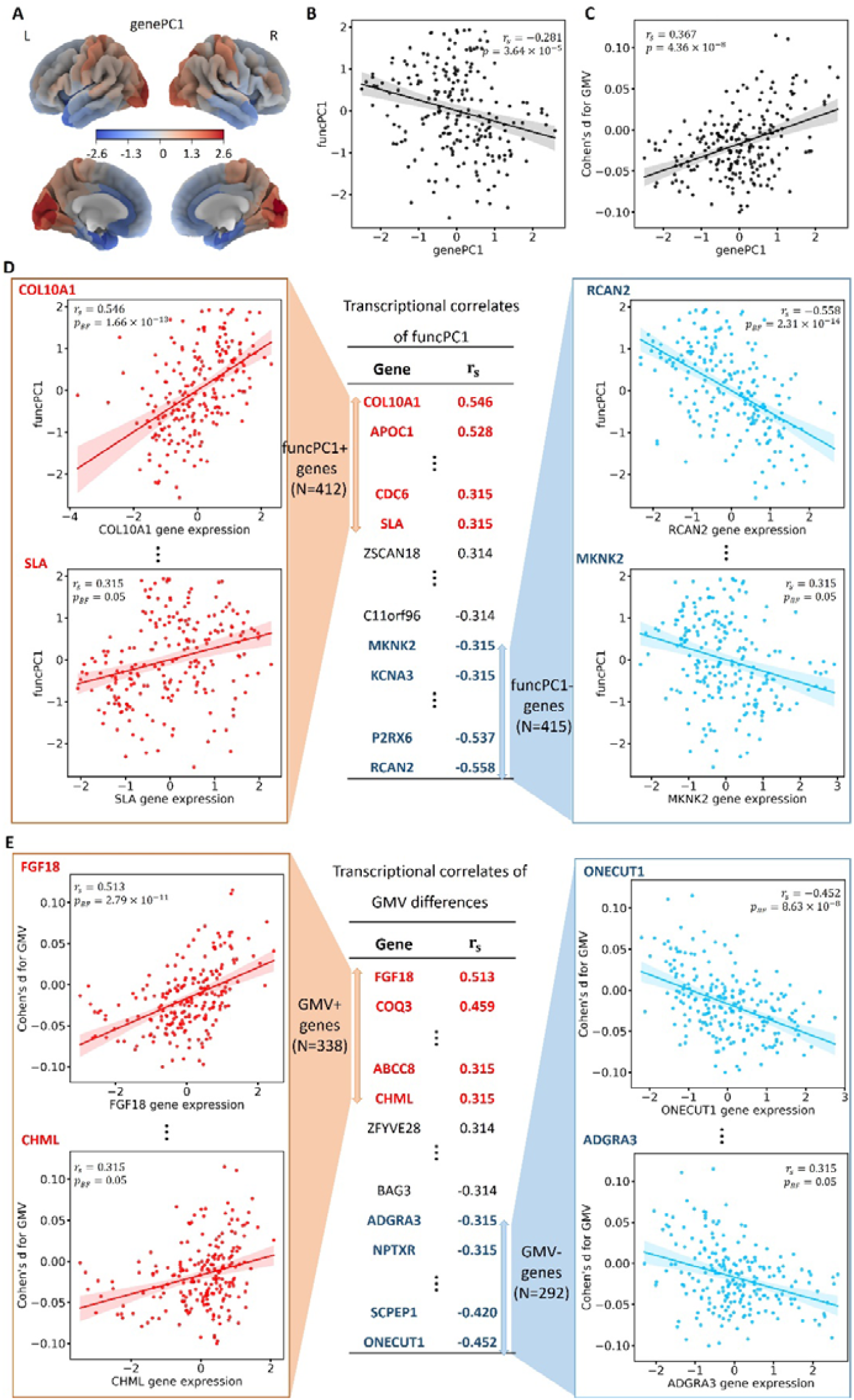
Relations of gene expression with case-control differences for 210 cortical regions. **(A)** The first principal component of gene expression (genePC1); **(B)** Relationships of genePC1 with the first principal component of three major functional measures (funcPC1); **(C)** Relationships of genePC1 with gray matter volume (GMV) differences; **(D)** Genes positively or negatively related to funcPC1 (funcPC1+ or funcPC1- genes); **(E)** Genes positively or negatively related to GMV differences (GMV+ or GMV- genes). *p*_BF_ indicates the p values corrected by Bonferroni multiple testing correction.

We then identified the transcriptional correlates of brain abnormalities in MDD. Expression on 210 cortical regions of each gene were related to the funcPC1 and GMV differences, respectively (**Tables S5-6**). After Bonferroni multiple testing correction, gene expression of 412 genes showed significant positive correlations and 415 genes showed significant negative correlations with funcPC1 (funcPC1+ genes and funcPC1- gene respectively), (**Figure 2D, Figure S8A**). It indicates that higher gene expression in cortical regions is associated with increased and decreased neuronal activity in MDD for funcPC1+ and funcPC1- genes, respectively. For GMV, we identified 338 positively and 298 negatively related genes (GMV+ genes and GMV- genes respectively), indicating identified genes overexpressed in cortical regions with increased and decreased GMV in MDD, respectively (**Figure 2E, Figure S8B**).

### Relationships of transcriptional correlates with gene dysregulation

We related in-vivo transcriptional correlates of brain abnormalities to post-mortem gene dysregulation in cortex of patients with MDD for genes. DGE values of genes for MDD were negatively correlated with their correlations with funcPC1, but not significantly correlated with transcriptional correlates of GMV differences (**Figure S9**). When genes were divided into 100 bins, we found a significant negative correlation with the transcriptional correlates of funcPC1, but a significant positive correlation with the transcriptional correlates of GMV differences (**Figure 3A-B**). The finding indicates that the transcriptional correlates of in vivo MDD-related brain abnormalities from healthy AHBA donors captured the information of differential gene expression in ex vivo brain tissue from MDD patients.

**Figure 3.**
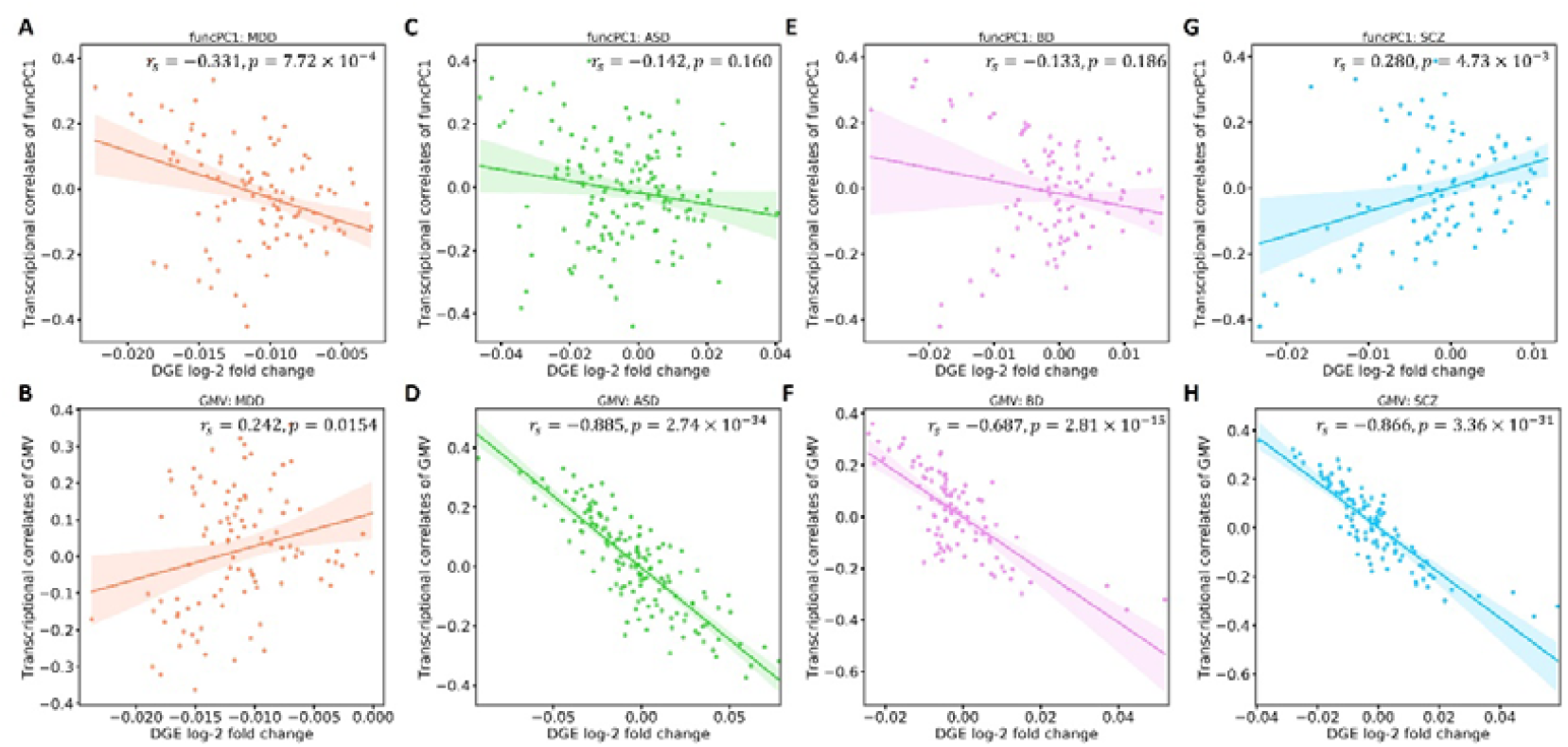
The correlations of the transcriptional correlates of brain abnormalities with differential gene expression (DGE) values for 100 gene bins. FuncPC1 is the first principal component of effect sizes for functional brain measures, and GMV indicates gray matter volume. Results are shown for major depressive disorder (MDD), autism spectrum disorder (ASD), bipolar disorder (BP), and schizophrenia (SCZ).

Through bin-based correlation analysis, the DGE values of ASD and BD also showed positive correlations with the transcriptional correlates of MDD-related GMV differences, but were not significantly associated with funcPC1 (**Figure 3C-F**). Paradoxically, DGE values of SCZ showed significant correlations in the opposite direction with a positive correlation to funcPC1, but a significant negative correlation to GMV differences (**Figure 3G-H**). This indicates that the transcriptional correlates of MDD-related brain abnormalities can capture the gene dysregulation of other psychiatric disorders. Comparable results were obtained at other bin numbers: 50, 150, and 200 gene bins (**Table S7**).

### Gene set enrichment

We found that all of our identified genes were significantly enriched for brain tissue, especially funcPC1+ and GMV- genes (**Figure 4A**). In tissue enrichment analysis, In cell enrichment analysis, the GMV- genes were specifically enriched for astrocytes, whereas the other three gene sets were significantly enriched for inhibitory and excitatory neurons (**Figure 4B**). Moreover, the funcPC1+ and funcPC1- genes also showed significant enrichment for microglia and endothelial cells, respectively.

**Figure 4.**
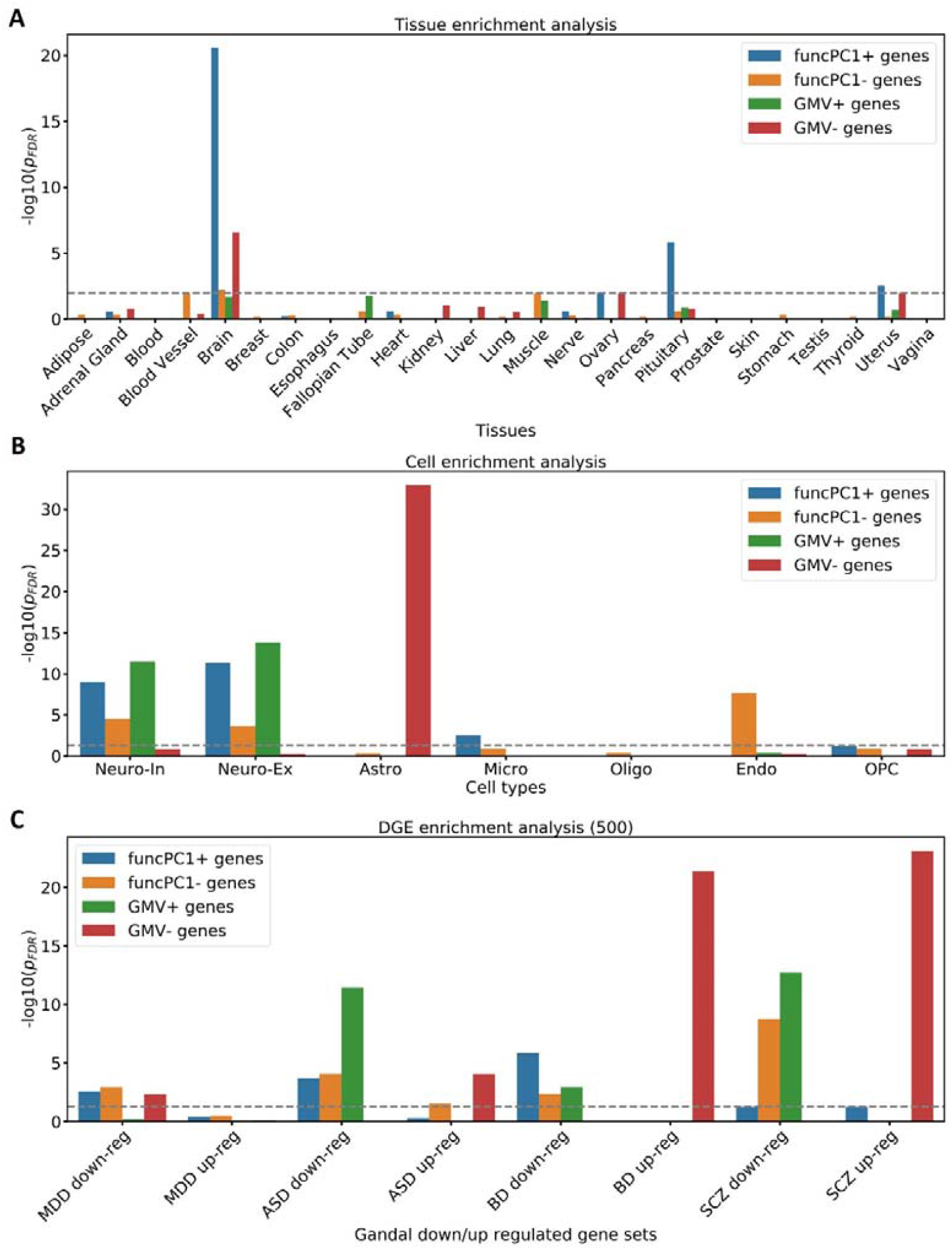
Enrichment analysis for tissues, cell types, and up- and down-regulated genes for psychiatric disorders. **(A)** Tissue specific expression analysis; **(B)** Cell enrichment analysis; **(C)** Enrichment analysis for genes up-regulated and down-regulated in psychiatric disorders. *p*_FDR_ is the adjusted *p* value after FDR multiple testing correction. FuncPC1 is the first principal component of effect sizes for functional brain measures, and GMV indicates gray matter volume. Genes positively and negatively related to funcPC1 and GMV differences are defined as funcPC1+, funcPC1- genes, GMV+, and GMV- genes. Neuro-In, inhibitory neuron; Neuro-Ex, excitatory neurons; Asto,astrocytes; Endo, endothelial; Mic, microglia; Oligo, oligodendrocytes; OPC, oligodendrocyte precursor cells. Major depressive disorder, MDD; autism spectrum disorder, ASD; bipolar disorder, BP; schizophrenia, SCZ.

We also selected up- and down-regulated genes for psychiatric disorders with the largest 500 positive and negative DGE values. We tested whether genes transcriptionally related to funcPC1 or GMV differences were enriched for genes that were highly up-regulated and down-regulated in psychiatric disorders (**Figure 4C**). The funcPC1+, funcPC1-, and GMV- genes were enriched for MDD down-regulated genes, but not for MDD up-regulated genes. In addition, widespread significant enrichments were identified for genes that were up- and down-regulated in ASD, BP, and SCZ. Of them, GMV- genes showed the strongest enrichment for up-regulated genes for BP and SCZ. Comparable results were observed using other thresholds (the largest 300-800 positive and negative DGE values) to define up- and down-regulated genes for psychiatric disorders(**Figure S10**).

We also conducted enrichment analysis for biological pathways including GO biological processes and KEGG pathways. The top 10 significant enrichment pathways are presented in **Figure 5**. For funcPC1+ genes, we observed enriched pathways such as synaptic transmission (i.e. modulation of chemical synaptic transmission, synaptic signaling, and synapse organization) and transport processes (i.e. regulation of ion transport, and negative regulation of transport) (**Figure 5A**). For funcPC1- genes, significant enrichment was mainly found for signaling pathways: Calcium signaling pathway, steroid hormone mediated signaling pathway, and enzyme-linked receptor protein signaling pathway (**Figure 5B**). In contrast, GMV- genes showed different enrichment terms, such as response to metal ion, neuroactive ligand-receptor interaction, and retrograde endocannabinoid signaling (**Figure 5C**), whereas GMV+ genes were only enriched for three pathways: inorganic ion transmembrane transport, sensory organ morphogenesis, and skeletal system development.

**Figure 5.**
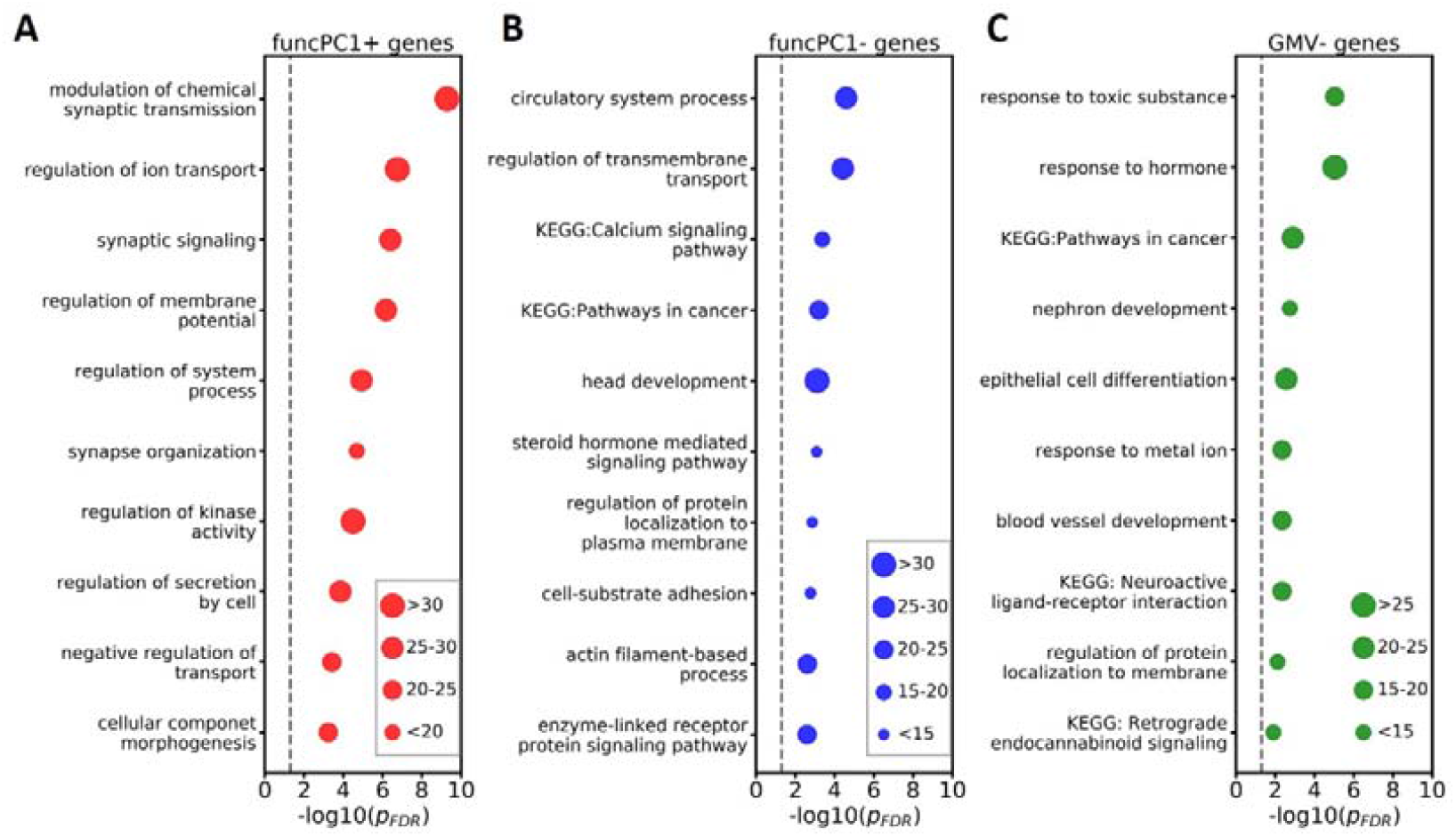
Top 10 enrichment pathways from the enrichment analysis for the gene ontology (GO) biological processes and Kyoto Encyclopedia of Genes and Genomes (KEGG) for different gene sets: **(A)** Genes positively related to the first component of effect sizes for functional measures (funcPC1+ genes); **(B)** Genes negatively related to the first component of effect sizes for functional measures (funcPC1- genes); **(C)** Genes negatively related to GMV differences (GMV- genes). *p*_FDR_ is the adjusted *p* value after FDR multiple testing correction.

## Discussion

In this study, we compared structural and functional brain abnormalities between MDD cases and controls. We observed similar case-control differences for ALFF, fALFF, and ReHo, and extracted the first principal component, funcPC1, which explained 71% variance of functional brain abnormalities in MDD, whereas smaller case-control differences were observed for GMV. We then related gene expression to funcPC1 and GMV differences, and found that whole gene expression (genePC1) was negatively associated with funcPC1, but positively associated with GMV differences. These opposite relationships were replicated in the UK Biobank, indicating that they exist in Asian and European populations. Moreover, we identified genes positively and negatively related to funcPC1 as well as GMV differences: funcPC1+, funcPC1-, GMV+, and GMV- genes. The identified genes were enriched for the brain tissue, cortical cells, and genes that were down-regulated in MDD. Furthermore, MDD-related genes showed different enrichment for biological pathways between brain function and structure. Together, these findings provide evidence for distinct genetic contributions to functional and structural brain abnormalities in MDD.

We found many functional abnormalities in MDD that were largely in line with previous studies [7, 8]. The abnormal regions, such as the anterior cingulate cortex and prefrontal cortex, are implicated in emotion regulation and cognitive functions, [37]. However, we did not observe significant differences in GMV. When relating gene expression to case-control differences in both the REST-meta-MDD and UK Biobank datasets, we found a negative correlation with brain function but a positive correlation with brain structure. This suggests that the gene expression patterns across the brain capture biologically meaningful and distinct information about MDD-related structural and functional brain abnormalities. A recent study also observed a positive correlation between gene expression and differences of morphometric similarity networks [25]. However, the study included a limited sample size (217 MDD patients and 208 healthy controls) and did not investigate brain function. Our results therefore extend these findings, and show that cortical gene expression can have divergent influences on structural versus functional brain abnormalities [38]. Another previous study combined structural and functional correlates of depression and negative affect across three datasets, but they ignored the potential differences in transcriptional correlates of brain structure and function. Moreover, they conducted statistical analysis in depressed individuals based on lifetime presence of depression or healthy adults with negative affect measures, so their findings do not reflect disease status at the time of MRI scanning [24]. Our results confirm that relations between gene expression and brain structure and function also occur during a depressive episode.

Through gene set enrichment analysis, we found that the identified genes showed specific expression in brain tissue and cortical cells, providing credibility for the observed transcriptome–neuroimaging relationships [17]. Interestingly, except for the GMV- genes, all gene sets were enriched for inhibitory and excitatory neurons, which have previously been related to depression [39]. For example, the somatostatin (SST) interneuron is one of the major inhibitory neuron types, which primarily target the apical dendrites of pyramidal neurons and provide synaptic and extrasynaptic inhibition [40]. Reduced dendritic inhibition from SST interneurons plays key roles in treatment-resistant depression [41]. Moreover, the gene expression by SST interneurons was significantly lower in postmortem cortical tissue from depressed individuals [42]. The parvalbumin (PV) neuron is another type of inhibitory neurons, which coordinates synchronous neuronal firing [43], and reductions in PV neurons in the prefrontal cortex were observed from postmortem MDD patients [44]. In addition, excitatory neurons were shown to be involved in reward circuits in depression [45]. For non-neurons, GMV- genes were significantly enriched in astrocytes, which influence synapse formation and elimination [46], and have been linked to MDD [47, 48].

The identified genes were enriched for genes that were down-regulated in MDD, and the transcriptional correlates of structural and functional brain abnormalities showed positive and negative correlations with gene dysregulation in postmortem tissue samples from MDD patients, respectively. This is in line with the identified relationships of whole gene expression with MDD-related differences in the cortex. Interestingly, the transcriptional correlates of MDD-related brain abnormities also showed significant associations with gene dysregulation in other psychiatric disorders. These findings could be in line with genetic pleiotropy, where a single gene influences multiple phenotypic traits [49], but could also be the result of a causal association, where certain abnormal gene expression in cortical regions could cause other psychiatric disorders, which may in turn cause depression.

The enrichment analysis was conducted for GO biological processes and KEGG pathways, and genes related to functional and structural abnormalities showed enriched biological pathways implicated in depression. For example, funcPC1+ genes were enriched for pathways related to synaptic transmission. Previous studies have proposed the hypothesis that the homeostatic mechanisms that control synaptic plasticity is disrupted in depression, resulting in destabilization and loss of synaptic connections in mood and emotion circuitry [50, 51]. The funcPC1- genes were enriched for signaling pathways, such as steroid hormone mediated signaling pathway. Previous research indicated that the depression may have a bidirectional association with altered steroid hormone secretion, leading to a vicious circle of more severe depression and more profound disturbances in steroid hormone signaling[52]. For GMV- genes, we observed enriched KEGG pathways of neuroactive ligand-receptor interaction and retrograde endocannabinoid signaling, which modulate various synaptic neurotransmissions or neural functions, including cognition, motor control, and pain [53, 54] and are related to MDD [55]. Overall, genes related to functional and structural abnormalities showed widespread and distinct enriched biological pathways.

While our study used a larger sample size, and first reported the distinct transcriptome-neuroimaging relationships between structural and functional abnormalities, there are also some limitations that need to be considered. First, AHBA expression data was measured based on tissues from six post-mortem adults with no known neuropsychiatric or neuropathological history. It was therefore not possible to examine the changes of regional expression levels of genes in depressed patients. Moreover, only two of six donors had samples in the left hemisphere, which may cause some bias for statistical analysis of the whole brain. Second, only GMV was used to measure brain structure in the REST-meta-MDD dataset, which only measures one aspect of brain structure. Nevertheless, we replicated this in the UK Biobank, in which we also observed a positive correlation of whole gene expression with case-control differences for cortical thickness. Third, although the relationships of whole gene expression with structural and functional abnormalities in MDD has been replicated in UK Biobank, the replication of transcriptional correlates for 15,633 individual genes was difficult to be implemented. This may be resulted by the biased definition of depressed individuals in UK Biobank dataset, which only considers two core symptoms of depression.

In summary, this study found opposite relationships of gene expression with structural and functional brain abnormalities in individuals with MDD. The transcriptional correlates of brain abnormalities showed significant correlations with gene dysregulation in postmortem tissue samples from MDD patients. Moreover, we identified genes involved in brain abnormalities, which were enriched for brain tissues, cortical, cells and biological pathways. These results highlight the importance of cortical gene expression for the development of brain abnormalities in MDD, and increase the understanding of the underlying neural mechanisms.

## Data Availability

All data produced in the present work are contained in the manuscript

## Acknowledgements

This research has been conducted using the REST-meta-MDD Project from the DIRECT Consortium and the UK Biobank Resource under Application Number 30091. A.A. & K.J.H.V. are supported by the Foundation Volksbond Rotterdam. This work was supported by a CSC grant to SL. GvW has received research funding by Philips for an unrelated project.

## References

1. Kessler RC, Berglund P, Demler O, Jin R, Koretz D, Merikangas KR, et al. The Epidemiology of Major Depressive Disorder: Results from the National Comorbidity Survey Replication (NCS-R). J Am Med Assoc. 2003.

2. James SL, Abate D, Abate KH, Abay SM, Abbafati C, Abbasi N, et al. Global, regional, and national incidence, prevalence, and years lived with disability for 354 Diseases and Injuries for 195 countries and territories, 1990-2017: A systematic analysis for the Global Burden of Disease Study 2017. Lancet. 2018. 2018. https://doi.org/10.1016/S0140-6736(18)32279-7.

3. Sullivan PF, Neale MC, Kendler KS. Genetic epidemiology of major depression: Review and meta-analysis. Am J Psychiatry. 2000.

4. Howard DM, Adams MJ, Clarke TK, Hafferty JD, Gibson J, Shirali M, et al. Genome-wide meta-analysis of depression identifies 102 independent variants and highlights the importance of the prefrontal brain regions. Nat Neurosci. 2019. 2019. https://doi.org/10.1038/s41593-018-0326-7.

5. Levinson DF, Mostafavi S, Milaneschi Y, Rivera M, Ripke S, Wray NR, et al. Genetic studies of major depressive disorder: Why are there no genome-wide association study findings and what can we do about it? Biol Psychiatry. 2014.

6. Grieve SM, Korgaonkar MS, Koslow SH, Gordon E, Williams LM. Widespread reductions in gray matter volume in depression. NeuroImage Clin. 2013. 2013. https://doi.org/10.1016/j.nicl.2013.08.016.

7. Kühn S, Gallinat J. Resting-state brain activity in schizophrenia and major depression: A quantitative meta-analysis. Schizophr Bull. 2013. 2013. https://doi.org/10.1093/schbul/sbr151.

8. Gray JP, Müller VI, Eickhoff SB, Fox PT. Multimodal abnormalities of brain structure and function in major depressive disorder: A meta-analysis of neuroimaging studies. Am J Psychiatry. 2020. 2020. https://doi.org/10.1176/appi.ajp.2019.19050560.

9. Javaheripour N, Li M, Chand T, Krug A, Kircher T, Dannlowski U, et al. Altered resting-state functional connectome in major depressive disorder: a mega-analysis from the PsyMRI consortium. Transl Psychiatry 2021 111. 2021;11:1–9.

10. Schmaal L, Hibar DP, Sämann PG, Hall GB, Baune BT, Jahanshad N, et al. Cortical abnormalities in adults and adolescents with major depression based on brain scans from 20 cohorts worldwide in the ENIGMA Major Depressive Disorder Working Group. Mol Psychiatry. 2017. 2017. https://doi.org/10.1038/mp.2016.60.

11. Scharinger C, Rabl U, Pezawas L, Kasper S. The genetic blueprint of major depressive disorder: Contributions of imaging genetics studies. World J Biol Psychiatry. 2011.

12. Schmitt S, Meller T, Stein F, Brosch K, Ringwald K, Pfarr JK, et al. Effects of polygenic risk for major mental disorders and cross-disorder on cortical complexity. Psychol Med. 2021:1–12.

13. D Y, B D, AJ F, SH W, R M, M R, et al. Polygenic risk for depression and the neural correlates of working memory in healthy subjects. Prog Neuropsychopharmacol Biol Psychiatry. 2017;79:67–76.

14. Naumova OY, Lee M, Rychkov SY, Vlasova N V, Grigorenko EL. Gene Expression in the Human Brain: The Current State of the Study of Specificity and Spatio-temporal Dynamics NIH Public Access. Child Dev. 2013;84:76–88.

15. Hawrylycz MJ, Lein ES, Guillozet-Bongaarts AL, Shen EH, Ng L, Miller JA, et al. An anatomically comprehensive atlas of the adult human brain transcriptome. Nature. 2012. 2012. https://doi.org/10.1038/nature11405.

16. ArnatkevicLiūtė A, Fulcher BD, Fornito A. A practical guide to linking brain-wide gene expression and neuroimaging data. Neuroimage. 2019;189:353–367.

17. Fu J, Liu F, Qin W, Xu Q, Yu C. Individual-Level Identification of Gene Expression Associated with Volume Differences among Neocortical Areas. Cereb Cortex. 2020. 2020. https://doi.org/10.1093/cercor/bhz333.

18. Ritchie J, Pantazatos SP, French L. Transcriptomic characterization of MRI contrast with focus on the T1-w/T2-w ratio in the cerebral cortex. Neuroimage. 2018. 2018. https://doi.org/10.1016/j.neuroimage.2018.03.027.

19. Richiardi J, Altmann A, Milazzo AC, Chang C, Chakravarty MM, Banaschewski T, et al. Correlated gene expression supports synchronous activity in brain networks. Science (80-). 2015. 2015. https://doi.org/10.1126/science.1255905.

20. Wang GZ, Belgard TG, Mao D, Chen L, Berto S, Preuss TM, et al. Correspondence between Resting-State Activity and Brain Gene Expression. Neuron. 2015. 2015. https://doi.org/10.1016/j.neuron.2015.10.022.

21. Romero-Garcia R, Warrier V, Bullmore ET, Baron-Cohen S, Bethlehem RAI. Synaptic and transcriptionally downregulated genes are associated with cortical thickness differences in autism. Mol Psychiatry. 2019. 2019. https://doi.org/10.1038/s41380-018-0023-7.

22. Romero-Garcia R, Seidlitz J, Whitaker KJ, Morgan SE, Jones PB, Goodyer IM, et al. Schizotypy-Related Magnetization of Cortex in Healthy Adolescence Is Colocated With Expression of Schizophrenia-Related Genes. Biol Psychiatry. 2020. 2020. https://doi.org/10.1016/j.biopsych.2019.12.005.

23. Morgan SE, Seidlitz J, Whitaker KJ, Romero-Garcia R, Clifton NE, Scarpazza C, et al. Cortical patterning of abnormal morphometric similarity in psychosis is associated with brain expression of schizophrenia-related genes. Proc Natl Acad Sci U S A. 2019. 2019. https://doi.org/10.1073/pnas.1820754116.

24. Anderson KM, Collins MA, Kong R, Fang K, Li J, He T, et al. Convergent molecular, cellular, and cortical neuroimaging signatures of major depressive disorder. Proc Natl Acad Sci U S A. 2020. 2020. https://doi.org/10.1073/pnas.2008004117.

25. Li J, Seidlitz J, Suckling J, Fan F, Ji GJ, Meng Y, et al. Cortical structural differences in major depressive disorder correlate with cell type-specific transcriptional signatures. Nat Commun. 2021. 2021. https://doi.org/10.1038/s41467-021-21943-5.

26. Fan L, Li H, Zhuo J, Zhang Y, Wang J, Chen L, et al. The Human Brainnetome Atlas: A New Brain Atlas Based on Connectional Architecture. Cereb Cortex. 2016. 2016. https://doi.org/10.1093/cercor/bhw157.

27. Yan CG, Chen X, Li L, Castellanos FX, Bai TJ, Bo QJ, et al. Reduced default mode network functional connectivity in patients with recurrent major depressive disorder. Proc Natl Acad Sci U S A. 2019. 2019. https://doi.org/10.1073/pnas.1900390116.

28. Chen X, Lu B, Li H-X, Li X-Y, Wang Y-W, et al. The DIRECT consortium and the REST-meta-MDD project: towards neuroimaging biomarkers of major depressive disorder. Psychoradiology. 2022;2:32–42.

29. Chao-Gan Y, Yu-Feng Z. DPARSF: A MATLAB toolbox for ‘pipeline’ data analysis of resting-state fMRI. Front Syst Neurosci. 2010. 2010. https://doi.org/10.3389/fnsys.2010.00013.

30. Friston KJ, Williams S, Howard R, Frackowiak RSJ, Turner R. Movement-related effects in fMRI time-series. Magn Reson Med. 1996;35:346–355.

31. Ashburner J. A fast diffeomorphic image registration algorithm. Neuroimage. 2007. 2007. https://doi.org/10.1016/j.neuroimage.2007.07.007.

32. Markello RD, Arnatkevičiūtė A, Poline JB, Fulcher BD, Fornito A, Misic B. Standardizing workflows in imaging transcriptomics with the Abagen toolbox. Elife. 2021. 2021. https://doi.org/10.7554/eLife.72129.

33. Gandal MJ, Haney JR, Parikshak NN, Leppa V, Ramaswami G, Hartl C, et al. Shared molecular neuropathology across major psychiatric disorders parallels polygenic overlap. Science (80-). 2018. 2018. https://doi.org/10.1126/science.aad6469.

34. Dougherty JD, Schmidt EF, Nakajima M, Heintz N. Analytical approaches to RNA profiling data for the identification of genes enriched in specific cells. Nucleic Acids Res. 2010. 2010. https://doi.org/10.1093/nar/gkq130.

35. Seidlitz J, Nadig A, Liu S, Bethlehem RAI, Vértes PE, Morgan SE, et al. Transcriptomic and cellular decoding of regional brain vulnerability to neurogenetic disorders. Nat Commun. 2020. 2020. https://doi.org/10.1038/s41467-020-17051-5.

36. Kuleshov M V., Jones MR, Rouillard AD, Fernandez NF, Duan Q, Wang Z, et al. Enrichr: a comprehensive gene set enrichment analysis web server 2016 update. Nucleic Acids Res. 2016. 2016. https://doi.org/10.1093/nar/gkw377.

37. Gazzola V, Spezio ML, Etzel JA, Castelli F, Adolphs R, Keysers C. Primary somatosensory cortex discriminates affective significance in social touch. Proc Natl Acad Sci U S A. 2012. 2012. https://doi.org/10.1073/pnas.1113211109.

38. Scheepens DS, van Waarde JA, Lok A, de Vries G, Denys DAJP, van Wingen GA. The link between structural and functional brain abnormalities in depression: A systematic review of multimodal neuroimaging studies. Front Psychiatry. 2020.

39. Persson J, Wall A, Weis J, Gingnell M, Antoni G, Lubberink M, et al. Inhibitory and excitatory neurotransmitter systems in depressed and healthy: A positron emission tomography and magnetic resonance spectroscopy study. Psychiatry Res - Neuroimaging. 2021. 2021 https://doi.org/10.1016/j.pscychresns.2021.111327.

40. Tremblay R, Lee S, Rudy B. GABAergic Interneurons in the Neocortex: From Cellular Properties to Circuits. Neuron. 2016.

41. Yao HK, Guet-McCreight A, Mazza F, Moradi Chameh H, Prevot TD, Griffiths JD, et al. Reduced inhibition in depression impairs stimulus processing in human cortical microcircuits. Cell Rep. 2022. 2022. https://doi.org/10.1016/j.celrep.2021.110232.

42. Seney ML, Tripp A, McCune S, A. Lewis D, Sibille E. Laminar and cellular analyses of reduced somatostatin gene expression in the subgenual anterior cingulate cortex in major depression. Neurobiol Dis. 2015. 2015. https://doi.org/10.1016/j.nbd.2014.10.005.

43. Perlman G, Tanti A, Mechawar N. Parvalbumin interneuron alterations in stress-related mood disorders: A systematic review. Neurobiol Stress. 2021.

44. Rajkowska G, O’Dwyer G, Teleki Z, Stockmeier CA, Miguel-Hidalgo JJ. GABAergic neurons immunoreactive for calcium binding proteins are reduced in the prefrontal cortex in major depression. Neuropsychopharmacology. 2007. 2007. https://doi.org/10.1038/sj.npp.1301234.

45. Thompson SM, Kallarackal AJ, Kvarta MD, Van Dyke AM, LeGates TA, Cai X. An excitatory synapse hypothesis of depression. Trends Neurosci. 2015.

46. Chung WS, Allen NJ, Eroglu C. Astrocytes control synapse formation, function, and elimination. Cold Spring Harb Perspect Biol. 2015. 2015. https://doi.org/10.1101/cshperspect.a020370.

47. Zhou X, Xiao Q, Xie L, Yang F, Wang L, Tu J. Astrocyte, a promising target for mood disorder interventions. Front Mol Neurosci. 2019.

48. Rajkowska G, Stockmeier C. Astrocyte Pathology in Major Depressive Disorder: Insights from Human Postmortem Brain Tissue. Curr Drug Targets. 2013. 2013. https://doi.org/10.2174/13894501113149990156.

49. Gratten J, Visscher PM. Genetic pleiotropy in complex traits and diseases: Implications for genomic medicine. Genome Med. 2016.

50. Duman RS, Aghajanian GK. Synaptic dysfunction in depression: Potential therapeutic targets. Science (80-). 2012.

51. Duman RS, Aghajanian GK, Sanacora G, Krystal JH. Synaptic plasticity and depression: New insights from stress and rapid-acting antidepressants. Nat Med. 2016.

52. Nowacki J, Wingenfeld K, Kaczmarczyk M, Chae WR, Salchow P, Abu-Tir I, et al. Steroid hormone secretion after stimulation of mineralocorticoid and NMDA receptors and cardiovascular risk in patients with depression. Transl Psychiatry. 2020. 2020. https://doi.org/10.1038/s41398-020-0789-7.

53. Castillo PE, Younts TJ, Chávez AE, Hashimotodani Y. Endocannabinoid Signaling and Synaptic Function. Neuron. 2012.

54. Fan T, Hu Y, Xin J, Zhao M, Wang J. Analyzing the genes and pathways related to major depressive disorder via a systems biology approach. Brain Behav. 2020. 2020. https://doi.org/10.1002/brb3.1502.

55. Mechoulam R, Parker LA. The endocannabinoid system and the brain. Annu Rev Psychol. 2013.

