## Supplementary Information for "Gene expression has distinct associations with brain structure and function in major depressive disorder"

### **Supplementary methods**

#### **Replication dataset (UK Biobank)**

We used UK Biobank data with brain scans as replication dataset [1]. All participants provided written informed consent and the project was granted ethical approval (reference: 11/NW/0382) by the National Health Service North West Centre for Research Ethics Committee. As shown in **Figure S2**, a total of 802 individuals were identified as depressed individuals using the following criteria: (1) Participants reported the presence of one of the core symptoms of depression as *frequency of depressed mood* (Data Field: 2050) or *unenthusiasm / disinterest* (Data Field: 2060) *in the last 2 weeks prior to brain scanning* that has been present “more than half the days” or “nearly every day”, and (2) Data for these participants were available for the following variables: T1 and resting-state functional MRI data as well as covariates including fluid intelligence score (Data Field: 20016), educational attainment (Data Field: 6138), sex (Data Field: 31), and age when attended assessment centre (Data Field: 21003). It should be noted that the two items about depression mentioned above were the only two symptoms of depression available in the UK Biobank that reflect the mood state during the MRI assessment [2, 3], whereas a total of five out of nine symptoms are required to get a full diagnosis of major depressive disorder (MDD). The controls had MRI data and covariates available, but reported no presence (“not at all”) of negative mood symptom or a loss of disinterest in the last 2 weeks prior to brain scanning. Moreover, the controls did not have a history of MDD, and never diagnosed to have any mental health problems. Of this control group we randomly selected 802 individuals matching age, sex, intelligence, and educational attainment with those of the depressed individuals (**Table S1**).

#### **Imaging procedures in UK Biobank**

As replication dataset, we used T1-weighted MRI and rsfMRI data from UK Biobank. The brain imaging scanners used were standard Siemens Skyra 3T running VD13A SP4 with a 32-channel head coil. T1 scanning lasted about 5 minutes with the following parameters: repetition time = 2000 ms; echo time = 2.1 ms; flip angle = 8°; matrix size = 256 × 256 mm; voxel size = 1 × 1 × 1 mm; number of slices = 208. The acquisition parameters for the rsfMRI data were TR = 735 ms, TE = 39 ms, flip angle = 52°, matrix size =88 × 88 mm, voxel size = 2.4 × 2.4 × 2.4 mm, number of slices = 64, and volumes = 490. A series of preprocessing procedures were applied for T1 and rsfMRI data (<http://biobank.ctsu.ox.ac.uk/crystal/crystal/docs/brain_mri.pdf>). UK Biobank provided cortical surface area (CSA) and cortical thickness (CT) of 66 regions based on the Desikan-Killiany (DK) atlas [4]. For rsfMRI data, we used the BRANT toolbox [5] to estimate the amplitude of low-frequency fluctuation (ALFF) and regional homogeneity (ReHo) to detect the regional intensity of spontaneous fluctuations in the BOLD signal, and extracted region values based on the DK atlas. High-pass temporal filtering has been applied to the rsfMRI data in UK Biobank which can cause calculation bias for fractional ALFF (fALFF), so we did not include fALFF in statistical analysis.

#### **Statistical analysis in UK Biobank**

We first used two-sample *t* tests to compare the neuroimaging measures for cortical thickness (CT), ALFF, and ReHo based on the DK atlas between depressed individuals and controls. The t-values were then converted to Cohen’s d effect sizes for ease of interpretation. The covariates, including age, sex, education, intelligence, and head motion, were regressed out of the brain measures. Principle component analysis (PCA) was used to extract the first components of effect sizes for ALFF and ReHo (funcPC1). Moreover, AHBA transcriptome gene expression data were mapped to the DK atlas, and the first component pf gene expression matrix (genePC1) was also extracted. To identify the global transcriptome-neuroimaging relationships, we related the genePC1 to funcPC1 and CT differences, respectively.

### **Supplementary Figures**

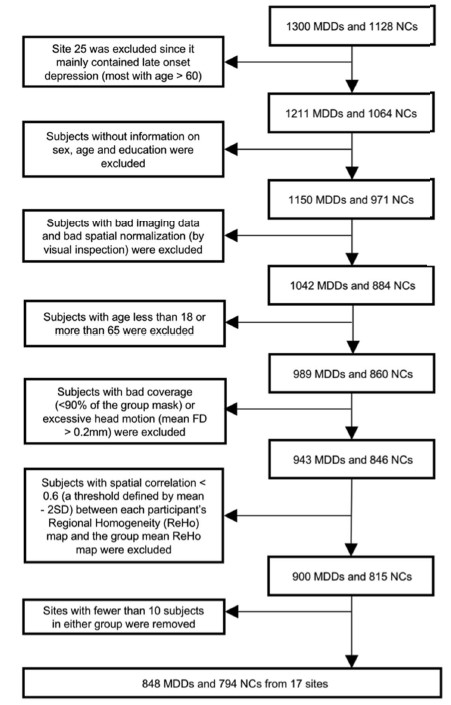

**Figure S1**. Sample selection of REST-meta-MDD dataset copied from the study by Yan et al. [6]. (The open access article has been distributed under Creative Commons Attribution-NonCommercial-NoDerivatives License 4.0 (CC BY-NC-ND).)

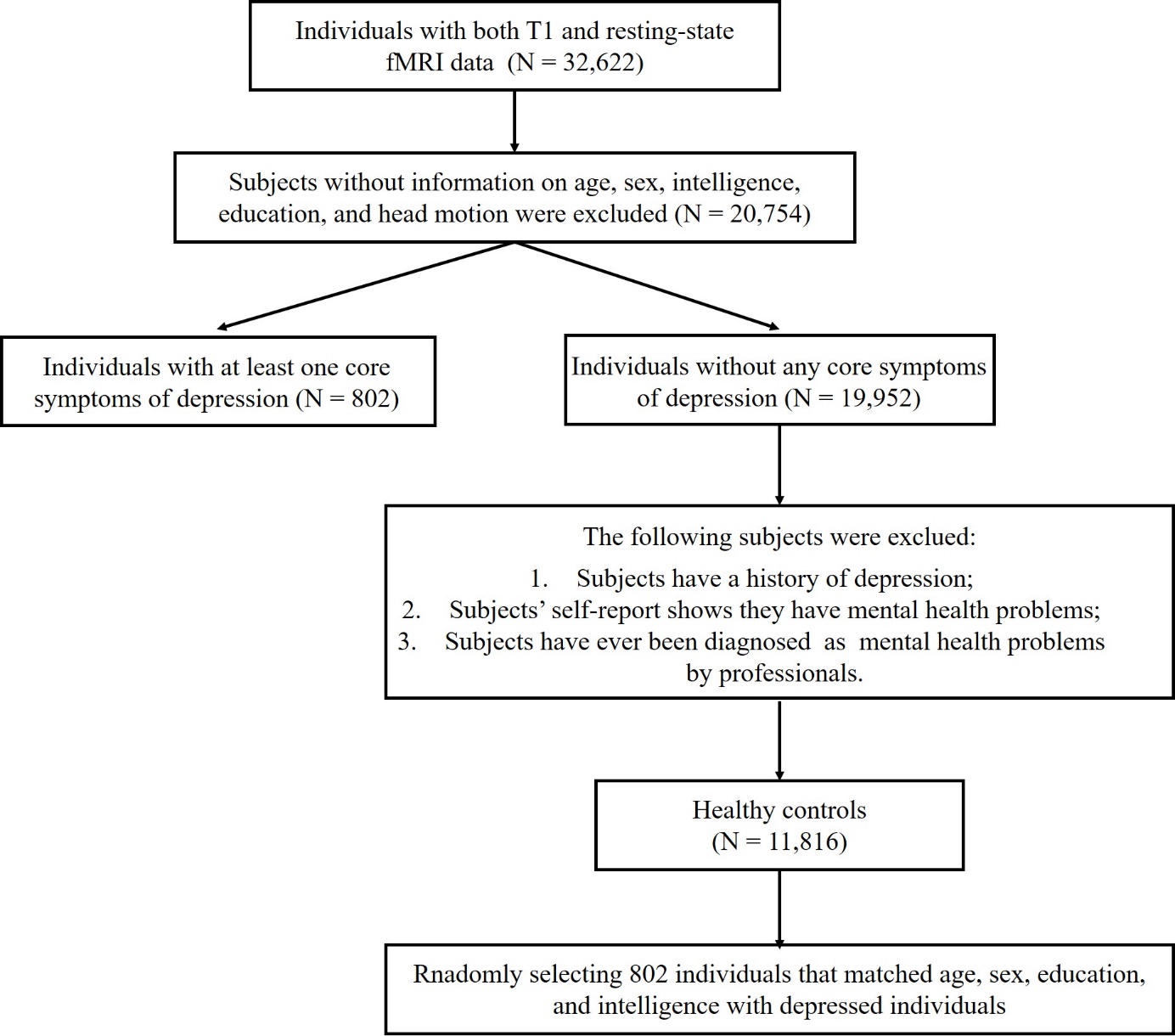

#### **Figure S2**. Sample selection of UK Biobank dataset.

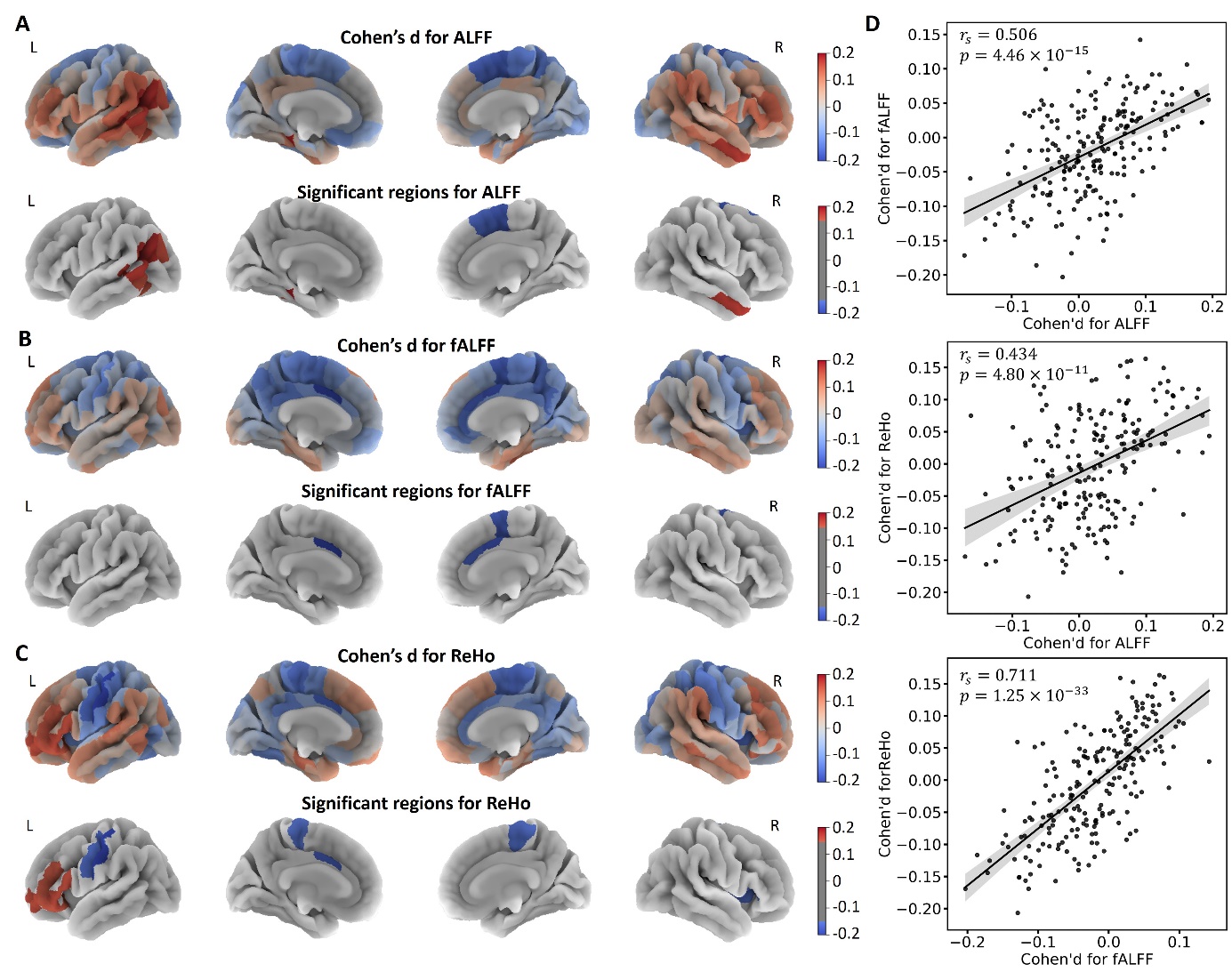

**Figure S3**. Case-control functional brain differences based on the Brainnetome (BN) atlas. **(A)** Differences (Cohen’s d) for amplitude of low-frequency fluctuation (ALFF), and statistically significant regions; **(B)** Differences (Cohen’s d) for fractional ALFF (fALFF), and statistically significant regions; **(C)** Differences (Cohen’s d) for regional homogeneity (ReHo) and statistically significant regions; **(D)** the correlations between effect sizes for functional brain measures.

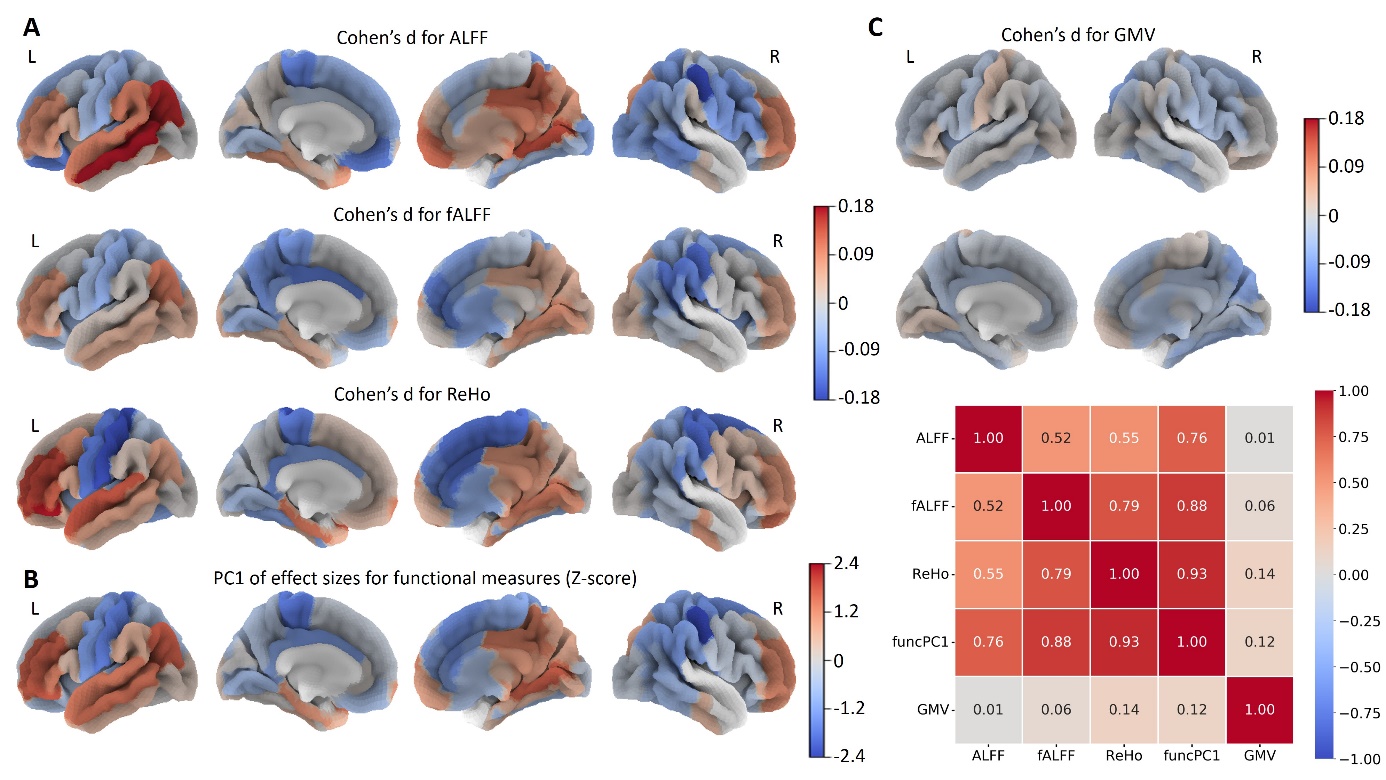

**Figure S4**. Case-control structural and functional brain differences in major depressive disorder (MDD) based on the Desikan-Killiany (DK) atlas. **(A)** Differences (Cohen’s d) for functional brain measures including amplitude of low-frequency fluctuation (ALFF), fractional ALFF (fALFF), and regional homogeneity (ReHo); **(B)** The first principle component of effect sizes for three functional measures (funcPC1) (Z-score normalization); **(C)** Differences (Cohen’s d) for gray matter volume (GMV); **(D)** The correlations between case-control differences.

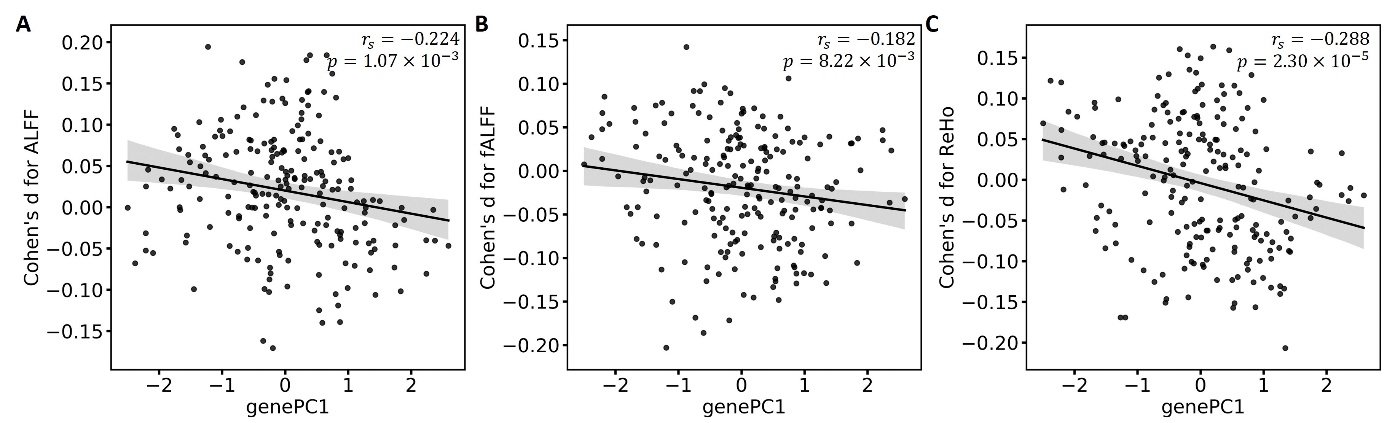

**Figure S5**. Relationships of gene expression with effect sizes for functional measures. **(A)** amplitude of low-frequency fluctuation (ALFF); **(B)** fractional ALFF (fALFF); **(C)** regional homogeneity (ReHo).

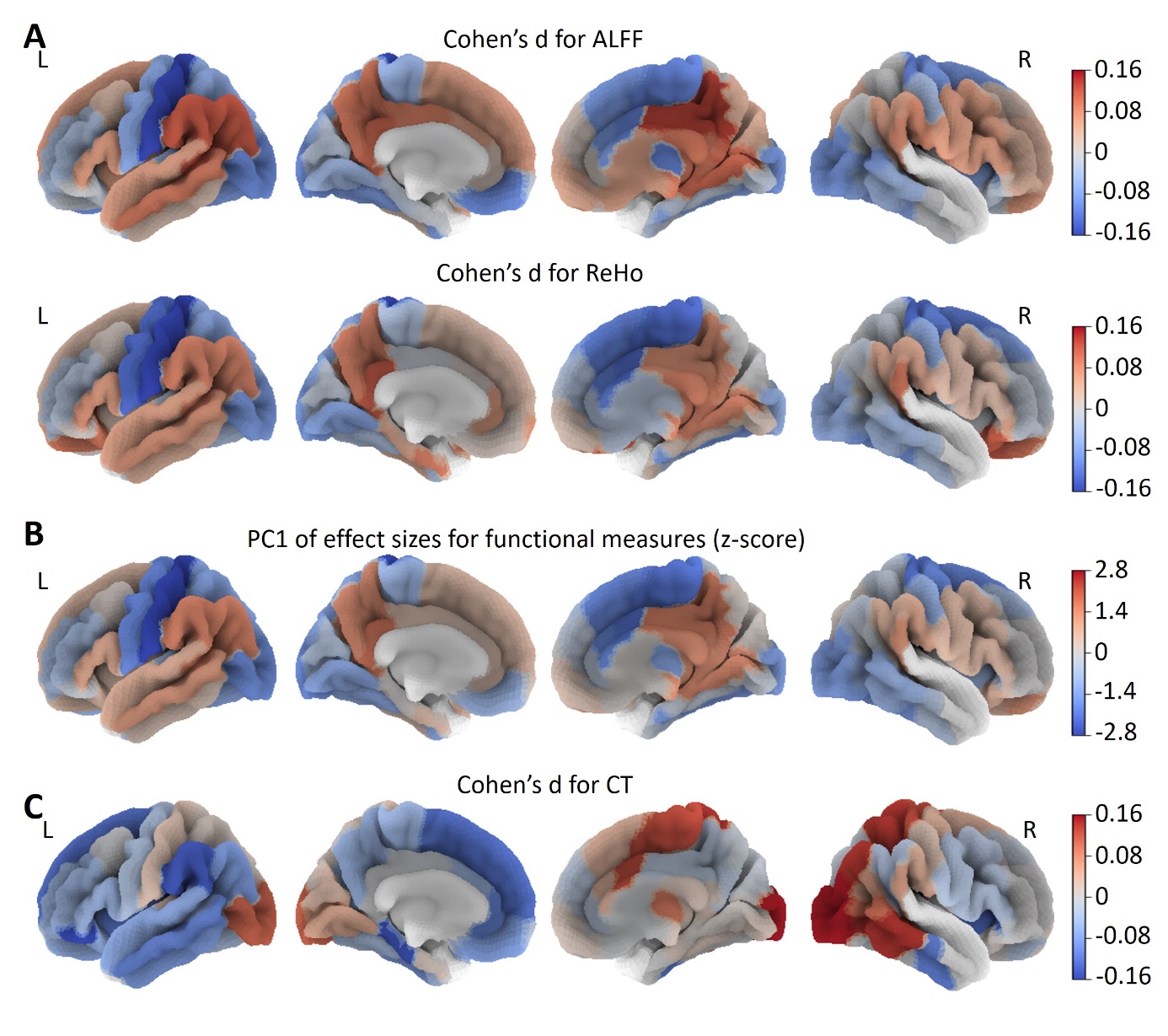

**Figure S6.** Case-control structural and functional brain differences in major depressive disorder (MDD) based on UK Biobank. **(A)** Differences (Cohen’s d) for functional brain measures including amplitude of low-frequency fluctuation (ALFF) and regional homogeneity (ReHo); **(B)** The first principle component of effect sizes for functional measures (funcPC1) (Z-score normalization); **(C)** Differences (Cohen’s d) for gray matter volume (GMV).

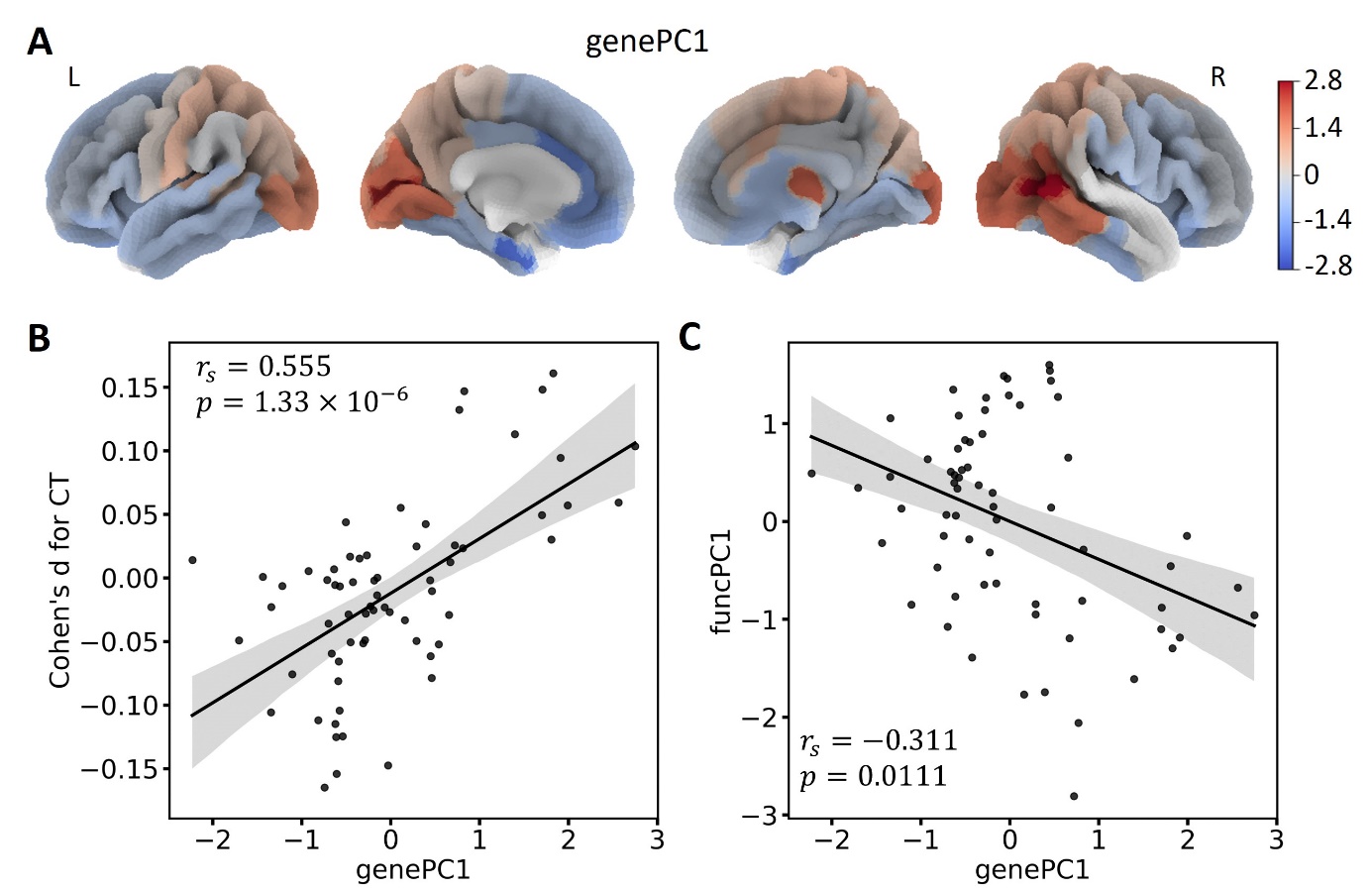

**Figure S7**. Relations of gene expression with case-control differences in UK Biobank. **(A)** The first principal component of gene expression (genePC1); **(B)** Relationships of genePC1 with cortical thickness (CT) differences; **(C)** Relationships of genePC1 with the first principal component of effect sizes for amplitude of low-frequency fluctuation (ALFF) and regional homogeneity (ReHo) (funcPC1).

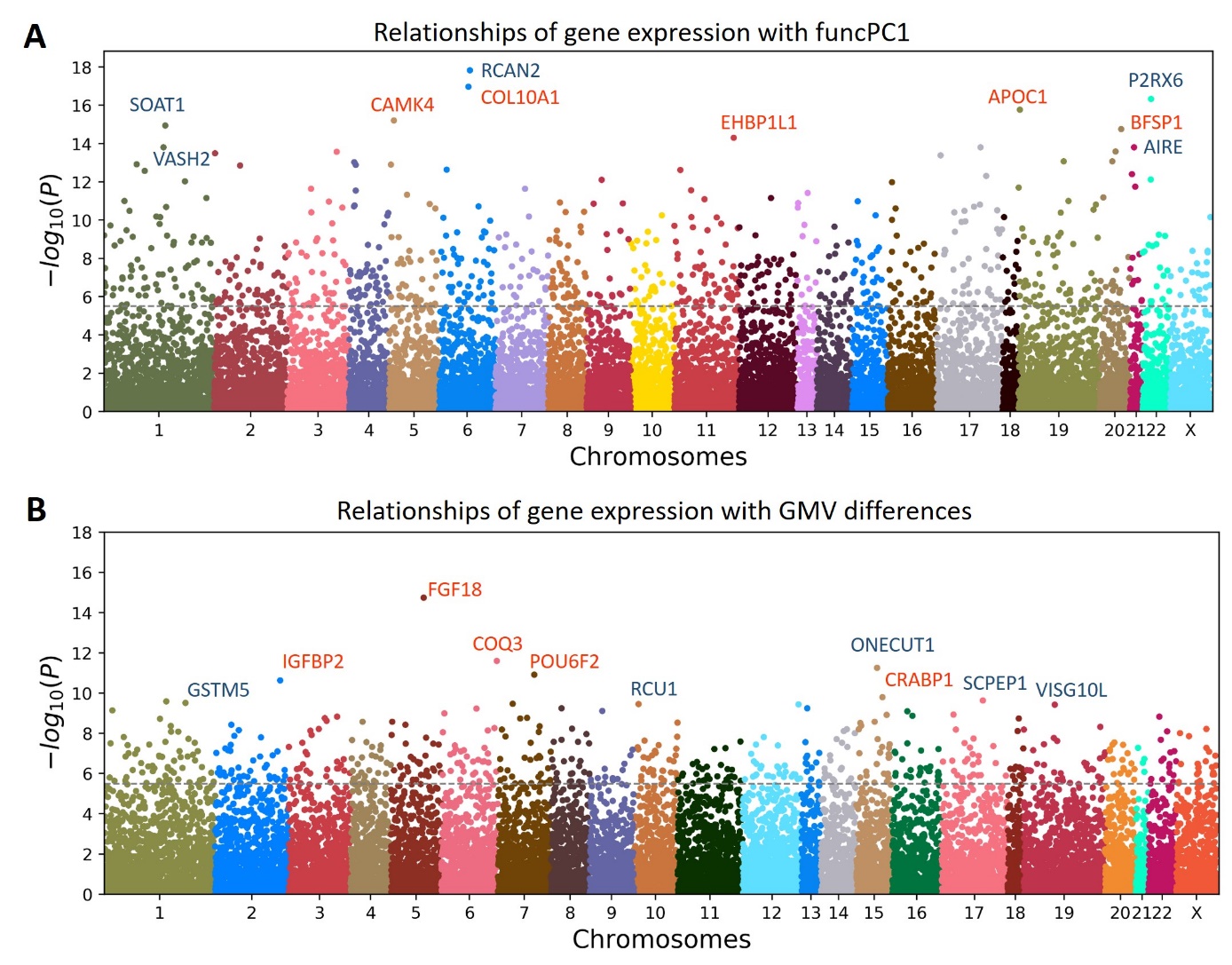

**Figure S8**. Manhattan plots showing associations between gene expression and cortical differences. **(A)** The first principal component of effect sizes for three major functional measures (funcPC1); **(B)** Gray matter volume (GMV) differences. X axis shows the chromosomes, and Y axis shows the –log_10_(p) value, which indicates the significance of the association of the gene expression with cortical differences. The horizontal dotted line indicates the significance threshold after correcting for multiple testing

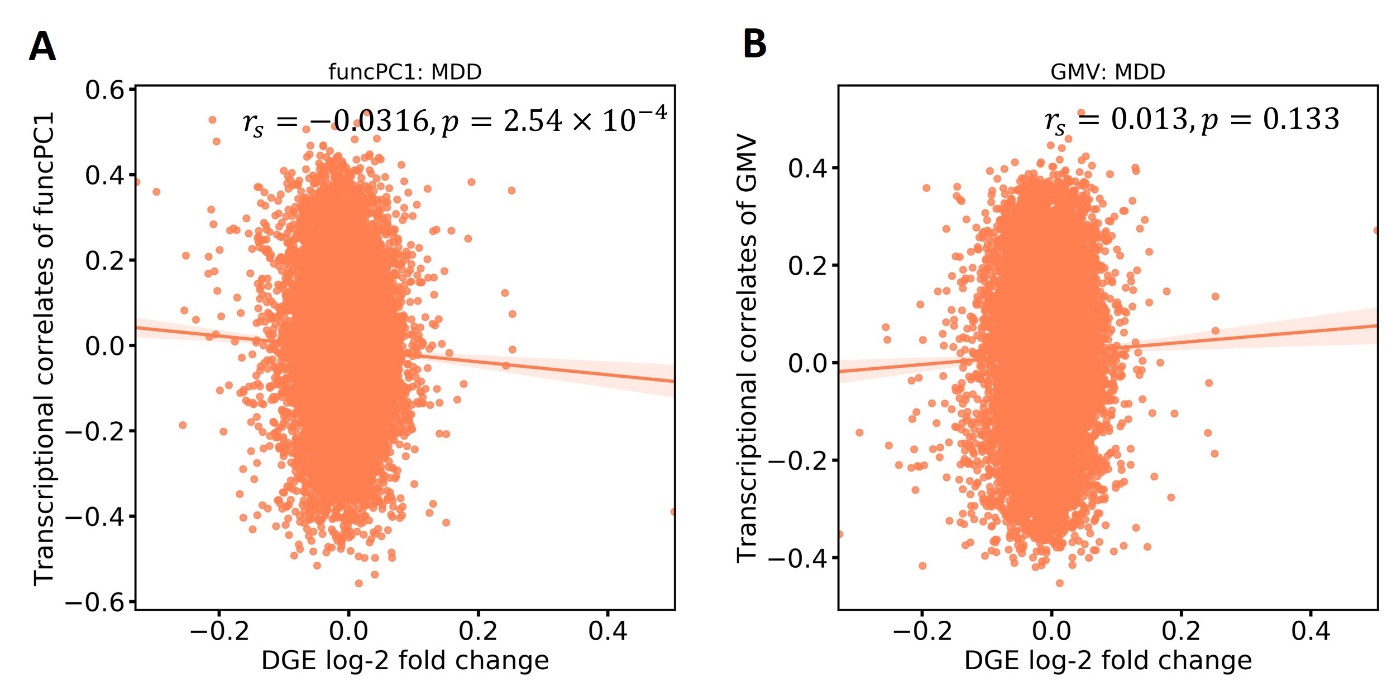

**Figure S9**. The correlations of the transcriptional correlates of brain abnormities with differential gene expression (DGE) values for major depressive disorder (MDD) when all genes are individually included. The funcPC1 is the first principal component of effect sizes for functional brain measures, and GMV indicates the gray matter volume.
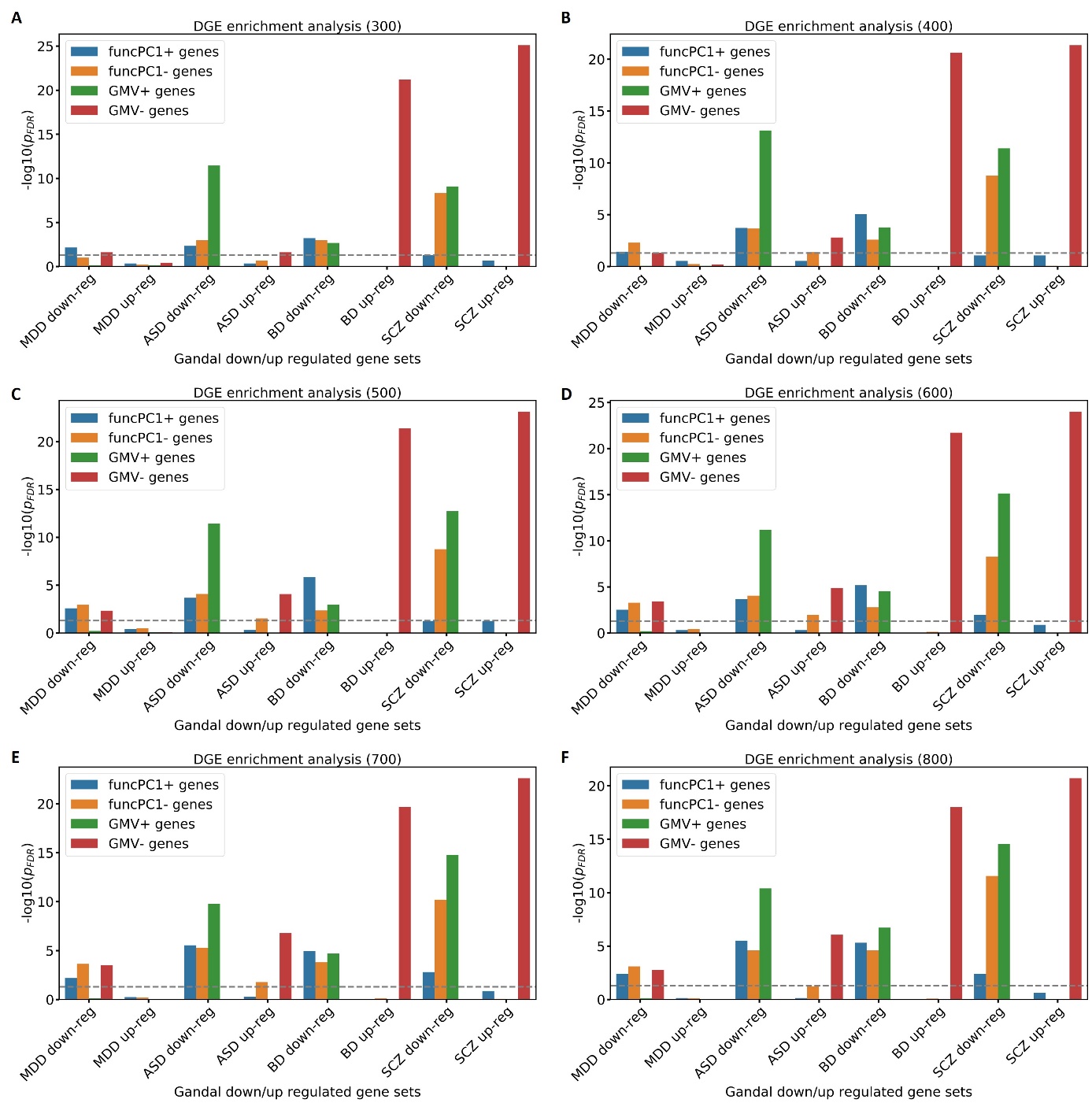

**Figure S10**. Enrichment analysis for up- and down-regulated genes at different thresholds (300-800) based on differential gene expression (DGE) analysis in psychiatric disorders. **(A)** 300; **(B)** 400; **(C)** 500; **(D)** 600; **(E)** 700; **(F)** 800. *p*_FDR_ is the adjusted *p* value after FDR multiple testing correction. FuncPC1 is the first principal component of effect sizes for functional brain measures, and GMV indicates the gray matter volume. Genes positively and negatively related to funcPC1 and GMV differences are defined as funcPC1+, funcPC1- genes, GMV+, and GMV- genes. Major depressive disorder (MDD), autism spectrum disorder (ASD), bipolar disorder (BP), and schizophrenia (SCZ).

### **References**

1.	Bycroft C, Freeman C, Petkova D, Band G, Elliott LT, Sharp K, et al. The UK Biobank resource with deep phenotyping and genomic data. Nature. 2018. 2018. https://doi.org/10.1038/s41586-018-0579-z.

2.	Smith DJ, Nicholl BI, Cullen B, Martin D, Ul-Haq Z, Evans J, et al. Prevalence and characteristics of probable major depression and bipolar disorder within UK Biobank: Cross-sectional study of 172,751 participants. PLoS One. 2013. 2013. https://doi.org/10.1371/journal.pone.0075362.

3.	Howard DM, Adams MJ, Shirali M, Clarke T-K, Marioni RE, Davies G, et al. Genome-wide association study of depression phenotypes in UK Biobank identifies variants in excitatory synaptic pathways. Nat Commun 2018 91. 2018;9:1–10.

4.	Desikan RS, Ségonne F, Fischl B, Quinn BT, Dickerson BC, Blacker D, et al. An automated labeling system for subdividing the human cerebral cortex on MRI scans into gyral based regions of interest. Neuroimage. 2006. 2006. https://doi.org/10.1016/j.neuroimage.2006.01.021.

5.	Xu K, Liu Y, Zhan Y, Ren J, Jiang T. BRANT: A Versatile and Extendable Resting-State fMRI Toolkit. Front Neuroinform. 2018. 2018. https://doi.org/10.3389/fninf.2018.00052.

6.	Yan CG, Chen X, Li L, Castellanos FX, Bai TJ, Bo QJ, et al. Reduced default mode network functional connectivity in patients with recurrent major depressive disorder. Proc Natl Acad Sci U S A. 2019. 2019. https://doi.org/10.1073/pnas.1900390116.
